# Relative effectiveness of non-surgical interventions for pain management in knee osteoarthritis: a protocol for a component network meta-analysis of randomised controlled trials

**DOI:** 10.1101/2020.12.22.20248700

**Authors:** Trevor Thompson, Bawan Ahmed, Sharon Weldon, Orestis Efthimiou, Brendon Stubbs

**Author notes:** Joint first authors (Trevor Thompson, Bawan Ahmed).

## Abstract

**Introduction:** Knee osteoarthritis is a chronic degenerative disease and the most common form of osteoarthritis, and is associated with significant chronic pain, disability and impairment of quality of life. Currently, there is no cure for knee osteoarthritis and pain management and improving quality of life are the main therapeutic goals. The objective of this study is to evaluate the relative efficacy and acceptability of currently available interventions using network meta-analysis in order to provide a comprehensive evidence base to guide future clinical treatment guidelines.

**Methods and analysis:** A comprehensive literature search of major electronic databases (MEDLINE, EMBASE, Cochrane Central Register of Controlled Trials) and clinical trial registries will be undertaken to identify randomised control trials (RCTs) of interventions listed in NICE guidelines for the treatment of knee osteoarthritis in adults. We will perform a network meta-analysis (NMA) to estimate relative intervention effects across the whole treatment network. If any studies use multicomponent interventions, we will employ a component network meta-analysis (CNMA) model to estimate the contribution of individual components. The quality of evidence will be assessed using the Confidence in Network Meta-Analysis (CINeMA) approach, which is based on the traditional GRADE framework adapted for NMA. Risk of bias will be assessed using the revised Cochrane Risk of Bias (RoB 2.0) tool for RCTs.

**Ethics and dissemination:** This study does not require ethical approval. Findings will be submitted to a peer-reviewed journal.

**PROSPERO registration number:** CRD42020184192.

**ARTICLE SUMMARY:** *Strengths and limitations of this study:*

- This will be the first network meta-analysis to assess relative effectiveness of interventions listed in NICE guidelines for pain management in knee osteoarthritis
- The study will provide an evidence base to inform future clinical guidelines and treatment decision making
- If relevant data are available, we will estimate the contribution of individual components in multicomponent interventions
- Quality of evidence underlying all treatments will be assessed
- Not all treatments will be evaluated if data are limited or we deem that network meta-analysis assumptions are violated

## 1 BACKGROUND

Osteoarthritis (OA) is a chronic degenerative musculoskeletal disorder of the joints, which most commonly affects the knee, hip, and hand [1]. OA is characterized by microscopic and macroscopic anatomical changes of the joint with a complex pathophysiological change of articular cartilage and subchondral bone [2]. This complex pathophysiology leads to cartilage degradation, bone remodelling, osteophyte formation and synovial inflammation, leading to pain, disability and loss of normal biomechanical joint function [3]. OA resulting from ‘wear and tear’ is by far the most common and typically develops at around 55-60 years, although secondary OA resulting from injury, congenital abnormality or inflammation can also occur [1].

OA is the most common form of arthritis and affects more than 300 million people worldwide [3]. Since OA is associated with major structural changes of the joints leading to pain and functional disability, its epidemiologic and economic burden on healthcare providers and society are substantial [4]. OA was estimated to be the fourth leading cause of disability by the year 2020 [5]. In the UK alone it is estimated that around 34% of people aged 45 years and older, approximately 8.75 million people, have sought treatment for OA [6]. The economic burden can be direct (e.g. treatments) or indirect (e.g. working days lost). A study in 2013 estimated that the negative economic impact of OA on UK economy was the equivalent of 1% GNP [7]. The most common form of OA affects the knee joint and accounts for almost 83% of OA burden [8, 9]. Furthermore, because of the knee joint’s anatomical location its impact on disability is substantial [10].

Currently there is no cure for OA. As pain is the most common symptom associated with OA, with more than 66% of patients with OA in constant pain [11], interventions are therefore primarily focused on pain management using a range of non-pharmacological and pharmacological interventions [3]. However, while there is an abundance of research evaluating the effectiveness of different interventions compared to placebo, the question of which treatment is optimal is inhibited by a lack of data on the relative efficacies of competing interventions. Similarly, a better understanding of the relative adverse effects of different treatments is needed, with the risk-to-benefit ratio a critical aspect of decision making. This is particularly true in the geriatric population in whom OA is most prevalent, and for whom treatment selection is often determined by adverse events, patient compliance and polypharmacy considerations [12].

Network meta-analysis (NMA) provides a powerful means of providing relative estimates of the effects of different interventions for a particular indication [13]. NMA allows the synthesis of data from both head-to-head trials and placebo-controlled trials, and can provide estimates of relative treatment effects among any two treatments in the network, even those that have not been previously compared in head-to-head trials. Furthermore, component NMA allows the estimation of the effects of individual components of combined interventions and can be especially useful for indications commonly treated by intervention packages [14].

The objective of this NMA is to assess the relative effectiveness and acceptability of treatments for the management of pain from knee osteoarthritis. As surgical interventions are generally used as a last resort, we will focus on non-surgical treatments and examine interventions listed in the NICE 2014 guidelines CG177 [15] or the NICE 2017 surveillance guideline update report [16]. The project is called Relative Effectiveness of Interventions for Knee Osteoarthritis (REIKO).

## 2 METHODS

This study protocol is registered on PROSPERO (registration number CRD42020184192). The protocol conforms to PRISMA-P guidelines [17] for the reporting of systematic review and meta-analysis protocols (see Supplementary File 1) and also incorporates additional considerations specific to NMA (Supplementary File 2). Eligibility criteria were developed using the PICOS framework, which are summarised in Table 1 and described in detail in the sections below.

**Table 1.**
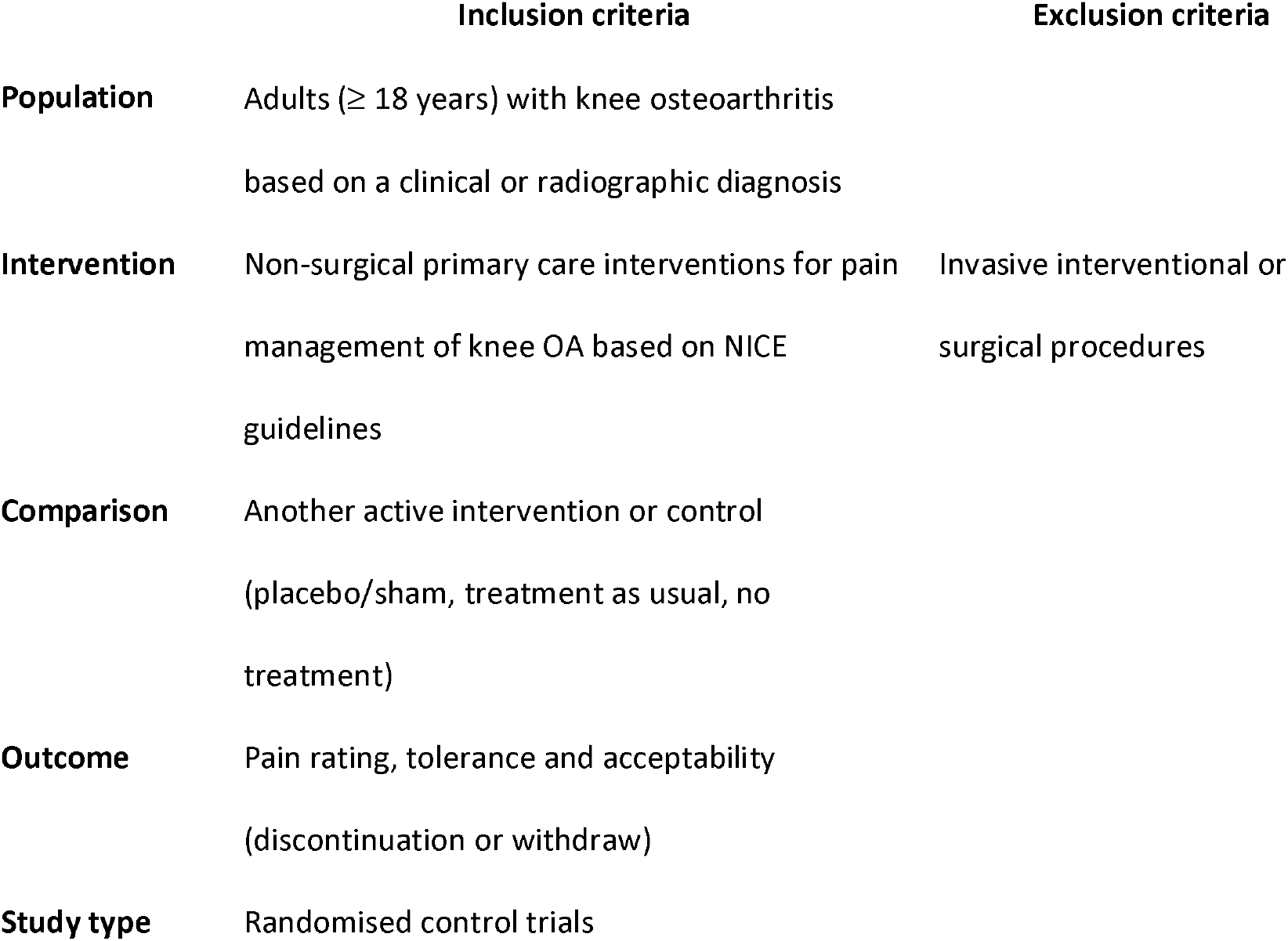
Brief summary of PICOS eligibility criteria (detailed descriptions in manuscript text)

### 2.1 Population

We will include studies of adults (≥ 18 years) with knee osteoarthritis based on a clinical or radiographic diagnosis. Where a mixed arthritis sample has been used, we will only include the study if (a) data for participants with knee osteoarthritis can be extracted separately, or (b) at least 75% of the sample has knee osteoarthritis.

### 2.2 Interventions

We will include interventions for pain management in knee osteoarthritis with any of the treatment components listed below. These interventions were chosen for inclusion as they are either recommended in NICE (2014) guidelines [CG177] for the management of knee osteoarthritis pain or listed in the NICE (2017) interim review as potential candidates for inclusion in future guidelines. The rationale for focusing on internationally recognised clinical guidelines is that these interventions more likely to be prescribed or recommended for patients.

#### Pharmacological

Paracetamol, topical NSAIDs, NSAIDs (non-selective) with or without a proton pump inhibitor, COX-2 inhibitors, opioids, topical capsaicin, duloxetine, intra-articular hyaluronan injections, intra-articular corticosteroid injection. *Non-pharmacological:* aerobic exercise, local muscle strengthening, education, local thermotherapy, TENS, electromagnetic field therapy, ultrasound.

Medications may be fixed or flexibly dosed. For medications approved for pain, we will only include study arms where dosages are within the licenced range. Where a drug is used off-label, we will include all trials but perform sensitivity analysis removing study arms using dosages outside the approved range for that drug’s indication. If different dosages have been used across multiple arms within a study, we will combine data and treat as a single node; unless multiple studies are available with similar dosing levels for that medication, in which case we will consider treating these as separate nodes.

### 2.3 Comparator(s)

A different eligible individual treatment or a control group (including placebo/sham, treatment as usual, no treatment).

### 2.4 Outcomes

#### 2.4.1 Primary outcome

Self-reported pain assessed with the following measures in order of prioritisation as recommended by Juhl et al. [18]: (i) the WOMAC pain subscale, (ii) a VAS/NRS pain rating measure, and (iii) any other acceptable instrument for pain listed in OARSI guidelines for the non-surgical management of osteoarthritis [19].

Assessment will be at the following distinct time periods following allocation to treatment: immediate (up to 2 weeks), short-term (closest to 3 months), medium term (closest to 6 months) and long-term follow-up (closest to 12 months). If these divisions fail to sensitively reflect the pattern of assessment timings used across studies, we may reclassify these windows prior to analysis to reflect trial practices.

As many pharmacological interventions are often trialled for immediate and short-term outcomes, and non-pharmacological treatment (e.g. exercise) trials may more frequently include long-term outcomes, separate analyses in each time window ensures that treatments are compared in time windows appropriate for how those interventions are used [20].

When pain ratings have been collected at multiple time points within a time window, we will use the time point closest to the median value across studies for the immediate and short-term windows and the longest follow-up for the long term follow-up window. If data were collected across multiple time points but only reported for a subset of these, we will make every possible attempt to retrieve all data to reduce the possibility of exaggerated treatment effects from selective reporting [21]. If we are unable to retrieve the preferred data, we will use outcomes at the next closest time point but conduct sensitivity analysis excluding these studies.

#### 2.4.2 Secondary outcomes

1. *Acceptability* –the proportion of patients in each group who withdraw before the end of the treatment for (a) any reason, and (b) due to adverse effects.
2. *Physical function* –any valid patient reported outcome of physical functioning, with prioritisation of the WOMAC function subscale.
3. *Stiffness*– any valid patient reported outcome of stiffness prioritising the WOMAC stiffness subscale.

### 2.5 Study Designs

Only randomised controlled trials (RCTs) comparing an active intervention with another eligible intervention or control will be included. Randomisation can be at the individual or group level. Standard errors from cluster RCTs will be adjusted to account for design effects using a standard formula [e.g., 22]. Both parallel group and crossover designs will be included, although for crossover designs we will extract data from the first trial period only to eliminate the possibility of carryover effects.

### 2.6 Measures of effect size

For continuous scores (pain, physical function, stiffness) we will compute the mean difference (MD) as the effect size, based on differences in post-treatment scores but using group differences in change scores if these are unavailable (which for randomised designs should give the same expected effect sizes as post-score differences, but often with larger variance).

As the WOMAC scale is available with three different response formats (0-4, 0-10 and 0-100) [23] we will normalise all WOMAC scores to the metric of the most commonly used format [24]. If a non-WOMAC measure have been used, we will attempt to normalise scores to the WOMAC metric using any conversion algorithm that may exist. If a study assesses pain both under load (e.g. while walking) and at rest we will take the average of these measures (consistent with the WOMAC scoring), otherwise we will use whichever of the two is reported and examine the impact of this decision in sensitivity analysis. If a substantial number of scale conversions are necessary, we will use the standardised mean differences as the effect size.

For the binary outcomes of acceptability, we will compute the odds ratio comparing the odds of discontinuation in one intervention with the odds of discontinuation in another intervention/control arm.

### 2.7 Information sources

The following bibliographic sources will be searched for studies indexed from database inception to the date of the search: MEDLINE, MEDLINE In-Process, EMBASE and the Cochrane Central Register of Controlled Trials. We will also search for unpublished and ongoing trials using the WHO International Clinical Trials Registry Platform (ICTRP) and ClinicalTrials.gov. It is important to include unpublished data, since the well-known bias towards publication of significant findings can, when relying on published literature alone, lead to an overestimation of treatment effects and an underestimation of adverse effects [25].

Where complete data for a relevant outcome are not available from a report of an eligible trial we will contact authors to request data. In addition, we will conduct a manual search of relevant reviews and reference lists of eligible studies.

### 2.8 Search strategy

A broad search strategy will be used to identify eligible trials using controlled vocabulary terms and free-text keywords of titles and abstracts relating to randomized trials, osteoarthritis and pain. For identifying randomized trials, we will use RCT filters provided by the Cochrane group for MEDLINE and by Wong et al. [26] for Embase. No publication date or language restrictions will be implemented, although studies for which a translation cannot be obtained will be listed as potentially eligible but not considered for full review. Searches will be re-run just before submission of the review and updated to account for any additional data. The search strategy for Medline (OVID) is provided in Supplementary File 3.

### 2.9 Study Selection

Titles and abstracts of each record returned by initial searches will be independently screened by BA and one other member of the review team, who will exclude studies not meeting eligibility criteria. BA and one other reviewer will then independently screen the full-text of remaining articles, retaining only eligible studies for inclusion in the network meta-analysis. Disagreements at any stage will be resolved through discussion or with a third member of the review team if necessary.

## 3 STATISTICAL ANALYSIS PLAN

### 3.1 Data extraction

One reviewer (BA) will perform data extraction and coding, with extracted data checked for accuracy by an experienced data analyst from the review team. We will use a standardized coding form based on our previous studies accompanied by an explanatory codebook. Information extracted will include: study characteristics (such as trial design, source of financial support, trial size, study location), participant and disease characteristics (such as mean age, male/female ratio, disease duration, disease severity and baseline pain severity), intervention and control details, outcome data (including timing of assessments and any information provided on missing data).

When available study data do not allow computation of effect sizes using standard formula we will: (a) extract inferential statistics (e.g. *F, p, t* etc) that allow effect sizes to be computed [e.g., 27], (b) contact study authors for data, (c) for missing SDs, used the pooled SD from other similar studies [28]. In the event of the same data published in multiple sources, we will extract data from the source which reports data with the most clarity. In the case of both published and unpublished data being available, we will prioritise published data which would have been subject to peer review, but we will conduct sensitivity analysis to examine the impact of this decision.

When a study reports participant drop-out, we will note whether effect sizes were computed from the reduced sample (i.e. per protocol) or the entire sample (i.e. intention-to-treat, e.g. using last observation carried forward) and prioritise intention-to-treat. When data are missing or ambiguously presented, we will contact study authors up to three times over a 6-week period for clarification.

### 3.2 Study characteristics and treatment network

We will provide a descriptive table summarising the key characteristics of each eligible study including interventions used, patient populations and trial characteristics. A network diagram will show which interventions were compared, with larger network nodes indicating a greater number of patients and thicker connecting lines between nodes indicating a greater number of trials.

### 3.3 Network meta-analysis (NMA)

Relative treatment effects across the whole network will be estimated with NMA. This method makes use of a wide pool of evidence by aggregating data from both direct head-to-head trials and indirect evidence (where two treatments can be compared indirectly via a common comparator such as placebo). The results of each treatment comparison will be provided in a Table, which will present results based on NMA estimates and on those from head-to-head trials only. The effectiveness of each treatment relative to a control reference will be presented in a forest plot. Mean ranks with their 95% credible intervals and SUCRA (a simple transformation of the mean rank) will be used to provide a hierarchy of the best treatments.

If combined intervention data are available, we will use component network meta-analysis (CNMA) to estimate the relative effects of different treatment components. CNMA [14] is able to estimate the effects of individual treatment components based on single component interventions (e.g. exercise only) and combined interventions (e.g. exercise + education) after disaggregation of the individual intervention components, and thus can provide greater statistical power than NMA with conventional parametrisation. Similarly, individual treatment component effects can be added to provide a more robust estimate of combined treatment effects compared to relying on direct combination studies only, and combinations can be modelled as additive or interactive (synergistic or antagonistic) effects [29]. This may be of particular benefit for osteoarthritis intervention research given that combination treatments can be a particularly effective way for older adults to manage their greater adverse safety profile from pharmacological agents [30] and that empirical data on combination treatments are relatively sparse [15].

Estimation will be based on a random-effects model using the *netmeta* package in R [31], with any additional analysis that cannot be performed in *netmeta* carried out in an alternative package (e.g. mvmeta in Stata).

#### 3.3.1 Transitivity and inconsistency in network meta-analysis

A fundamental assumption of NMA in general is that studies of different treatment comparisons should be similar in effect modifiers (participant and study characteristics that modify the relative efficacy of the treatments). To help assess this assumption, we will create and visually inspect a table summary of the distribution of potential effect modifiers (e.g. baseline pain severity, OA severity, age, gender) across different treatment comparisons. We will also compare direct and indirect estimates for consistency, both globally across the whole network using a design by treatment interaction approach [32] and locally for each comparison using the ‘back-calculation’ method [33]. Using the latter method we will highlight any substantive difference between direct and indirect estimates and advise caution in the interpretation of the corresponding NMA estimates.

We will proceed with NMA in the case of minor dissimilarities, and if there are sufficient data, we will explore the influence of effect modifiers on inconsistency using network meta-regression. In the event of considerable dissimilarity, we will consider excluding network nodes or not proceeding with NMA if substantive inconsistency across nodes is still present.

If there is evidence of sequestration of pharmacological and non-pharmacological treatments, such that there are no or very limited head-to-head pharmacological vs. non-pharmacological comparisons, we will split the network and conduct separate analyses on pharmacological and non-pharmacological treatment networks separately and we will assume a common heterogeneity variance τ^2^ for each network. If splitting the network is not required, we will use one common heterogeneity variance for each of medication interventions vs. control, non-pharmaceutical interventions vs. control and medication vs. non-pharmaceutical comparisons if data suggests τ^2^ is notably different for these three networks [34, 35].

#### 3.3.2 Additivity assumption in component network meta-analysis

An assumption of CNMA is that summing individual treatment component effects to estimate the effect of a combined treatment reliably reflects the effect of this combination administered in practice [29]. To help determine the plausibility of this assumption, we will check for consistency between effects estimated for combination interventions based on the additive model with those from any direct combination studies [36]. We will also carry out a test of additivity [29], based on a comparison of treatment estimates from the standard NMA model and the additive CNMA model. We will use clinical judgement to assess whether it is plausible that the components involved in a treatment combination are likely to operate through different pathways [36] and therefore whether considering effects to be additive is appropriate. We will also compare the fit of additive and interaction models with the Deviance Information Criteria statistic, and favour the additive model unless the interaction model is a significantly better fit. If any of these checks suggests intractable problems we will conduct only a standard NMA where combination treatments are entered as distinct nodes in the network rather than estimating these based on a component NMA model.

### 3.4 Meta-regression and sensitivity analysis

If there are sufficient data, we will conduct network meta-regression to identify possible sources of notable heterogeneity adding the effect modifiers listed previously. We will also use meta-regression to examine whether relative treatment effects are moderated by the following: diagnostic method (clinical/radiographic), dosage (above or below the median dosage) and industry sponsorship (yes/no).

We will also assess the robustness of the findings to various decisions by performing sensitivity analyses removing studies with high risk of bias, samples with secondary OA and where scale conversions have been performed.

### 3.5 Publication bias

We will visually examine contour enhanced treatment-control comparison adjusted funnel plots when ten or more studies are available, and further explore small study effects with Egger’s test for continuous outcomes and Peters’ test for binary outcomes [37, 38]. If bias is suspected we will explore this by including sample size as a covariate.

### 3.6 Evidence grading

The quality of the study evidence for the primary outcome of pain will be evaluated using the Confidence in Network Meta-Analysis (CINeMA) and presented in a Summary of Findings table. This approach was developed by Salanti et al. [39] and recently refined by Papakonstantinou et al. [40]. CINeMA is based on the traditional GRADE framework adapted for NMA, and provides a single overall confidence rating (‘high’, ‘moderate’, ‘low’, ‘very low’) for each pairwise comparison based on the six domains: (i) within-study bias, (ii) reporting bias, (iii) indirectness, (iv) imprecision, (v) heterogeneity, and (vi) incoherence. The overall confidence rating for each comparison is based on individual study ratings applying weights that mimic the study’s contribution to the treatment effect size.

Within-study bias will be assessed with the revised Cochrane Risk of Bias (RoB 2.0) tool for RCTs [41], which rates potential for study bias arising from the randomization process, deviations from the intended intervention, missing outcome data, measurement of outcomes and selective reporting. Risk of bias and evidence appraisal will be conducted independently by two reviewers, with any disagreement resolved by discussion or arbitration by a third reviewer if required. We will present network plots [40] to illustrate the level of bias in each comparison.

## Supporting information

Supplementary File 1. PRISMA-P-checklist

Supplementary File 2. PRISMA NMA checklist

Supplementary File 3. Medline search string

## Data Availability

n/a (study protocol)

## PATIENT AND PUBLIC INVOLVEMENT

This study is a synthesis of secondary data and will not require patient or public involvement.

## ETHICS AND DISSEMINATION

This study does not require ethical approval. We will disseminate our findings by publishing results in a peer-reviewed journal.

## AUTHOR CONTRIBUTIONS

TT and BA contributed equally to this paper. TT conceived the study idea and design and wrote the statistical analysis plan, and BA wrote the majority of the remainder of the protocol. All authors evaluated the study protocol critically for important intellectual content and contributed to critical revisions. All the authors gave their approval to the final protocol.

