## Supplementary File 2. PRISMA NMA checklist for "Relative effectiveness of non-surgical interventions for pain management in knee osteoarthritis: a protocol for a component network meta-analysis of randomised controlled trials"

**PRISMA NMA Checklist of Items to Include When Reporting A Systematic Review Involving a Network Meta-analysis**

| Section/Topic | Item # | Checklist Item | Reported on Page # |
|---|---|---|---|
| **TITLE** | | | |
| Title | 1 | Identify the report as a systematic review *incorporating a network meta-analysis (or related form of meta-analysis).* | 1 |
| **ABSTRACT** | | | |
| Structured summary | 2 | Provide a structured summary including, as applicable:<br>**Background:** main objectives<br>**Methods:** data sources; study eligibility criteria, participants, and interventions; study appraisal; and *synthesis methods, such as network meta-analysis.*<br>**Results**: number of studies and participants identified; summary estimates with corresponding confidence/credible intervals; *treatment rankings may also be discussed. Authors may choose to summarize pairwise comparisons against a chosen treatment included in their analyses for brevity.*<br>**Discussion/Conclusions:** limitations; conclusions and implications of findings.<br>**Other:** primary source of funding; systematic review registration number with registry name. | 2 |
| **INTRODUCTION** | | | |
| Rationale | 3 | Describe the rationale for the review in the context of what is already known*, including mention of why a network meta-analysis has been conducted.* | 5 |
| Objectives | 4 | Provide an explicit statement of questions being addressed, with reference to participants, interventions, comparisons, outcomes, and study design (PICOS). | 5-6 |
| **METHODS** | | | |
| Protocol and registration | 5 | Indicate whether a review protocol exists and if and where it can be accessed (e.g., Web address); and, if available, provide registration information, including registration number. | 6 |
| Eligibility criteria | 6 | Specify study characteristics (e.g., PICOS, length of follow-up) and report characteristics (e.g., years considered, language, publication status) used as criteria for eligibility, giving rationale. *Clearly describe eligible treatments included in the treatment network, and note whether any have been clustered or merged into the same node (with justification).* | 6-9 |

| | | | |
|---|---|---|---|
| Information sources | 7 | Describe all information sources (e.g., databases with dates of coverage, contact with study authors to identify additional studies) in the search and date last searched. | 11 |
| Search | 8 | Present full electronic search strategy for at least one database, including any limits used, such that it could be repeated. | |
| Study selection | 9 | State the process for selecting studies (i.e., screening, eligibility, included in systematic review, and, if applicable, included in the meta-analysis). | 12 |
| Data collection process | 10 | Describe method of data extraction from reports (e.g., piloted forms, independently, in duplicate) and any processes for obtaining and confirming data from investigators. | 12-13 |
| Data items | 11 | List and define all variables for which data were sought (e.g., PICOS, funding sources) and any assumptions and simplifications made. | 12-13 |
| **Geometry of the network** | **S1** | Describe methods used to explore the geometry of the treatment network under study and potential biases related to it. This should include how the evidence base has been graphically summarized for presentation, and what characteristics were compiled and used to describe the evidence base to readers. | 13 |
| Risk of bias within individual studies | 12 | Describe methods used for assessing risk of bias of individual studies (including specification of whether this was done at the study or outcome level), and how this information is to be used in any data synthesis. | 17-18 |
| Summary measures | 13 | State the principal summary measures (e.g., risk ratio, difference in means). *Also describe the use of additional summary measures assessed, such as treatment rankings and surface under the cumulative ranking curve (SUCRA) values, as well as modified approaches used to present summary findings from meta-analyses.* | 10, 13 |
| Planned methods of analysis | 14 | Describe the methods of handling data and combining results of studies for each network meta-analysis. This should include, but not be limited to:<br>• *Handling of multi-arm trials;*<br>• *Selection of variance structure;*<br>• *Selection of prior distributions in Bayesian analyses; and*<br>• *Assessment of model fit.* | 12-16 |
| **Assessment of Inconsistency** | **S2** | Describe the statistical methods used to evaluate the agreement of direct and indirect evidence in the treatment network(s) studied. Describe efforts taken to address its presence when found. | 14-15 |
| Risk of bias across studies | 15 | Specify any assessment of risk of bias that may affect the cumulative evidence (e.g., publication bias, selective reporting within studies). | 17-18 |
| Additional analyses | 16 | Describe methods of additional analyses if done, indicating which were pre-specified. This may include, but not be limited to, the following:<br>• Sensitivity or subgroup analyses;<br>• Meta-regression analyses;<br>• *Alternative formulations of the treatment network; and* | 13-16 |

| | | |
|---|---|---|
| | | • *Use of alternative prior distributions for Bayesian analyses (if applicable).* |

**RESULTS†**

| | | | |
|---|---|---|---|
| Study selection | 17 | Give numbers of studies screened, assessed for eligibility, and included in the review, with reasons for exclusions at each stage, ideally with a flow diagram. | NA |
| **Presentation of network structure** | **S3** | Provide a network graph of the included studies to enable visualization of the geometry of the treatment network. | NA |
| **Summary of network geometry** | **S4** | Provide a brief overview of characteristics of the treatment network. This may include commentary on the abundance of trials and randomized patients for the different interventions and pairwise comparisons in the network, gaps of evidence in the treatment network, and potential biases reflected by the network structure. | NA |
| Study characteristics | 18 | For each study, present characteristics for which data were extracted (e.g., study size, PICOS, follow-up period) and provide the citations. | NA |
| Risk of bias within studies | 19 | Present data on risk of bias of each study and, if available, any outcome level assessment. | NA |
| Results of individual studies | 20 | For all outcomes considered (benefits or harms), present, for each study: 1) simple summary data for each intervention group, and 2) effect estimates and confidence intervals. *Modified approaches may be needed to deal with information from larger networks.* | NA |
| Synthesis of results | 21 | Present results of each meta-analysis done, including confidence/credible intervals. *In larger networks, authors may focus on comparisons versus a particular comparator (e.g. placebo or standard care), with full findings presented in an appendix. League tables and forest plots may be considered to summarize pairwise comparisons.* If additional summary measures were explored (such as treatment rankings), these should also be presented. | NA |
| **Exploration for inconsistency** | **S5** | Describe results from investigations of inconsistency. This may include such information as measures of model fit to compare consistency and inconsistency models, *P* values from statistical tests, or summary of inconsistency estimates from different parts of the treatment network. | NA |
| Risk of bias across studies | 22 | Present results of any assessment of risk of bias across studies for the evidence base being studied. | NA |
| Results of additional analyses | 23 | Give results of additional analyses, if done (e.g., sensitivity or subgroup analyses, meta-regression analyses, *alternative network geometries studied, alternative choice of prior distributions for Bayesian analyses,* and so forth). | NA |

**DISCUSSION**

| | | | |
|---|---|---|---|
| Summary of evidence | 24 | Summarize the main findings, including the strength of evidence for each main outcome; consider their relevance to key groups (e.g., healthcare providers, users, and policy-makers). | NA |
| Limitations | 25 | Discuss limitations at study and outcome level (e.g., risk of bias), and at review level (e.g., incomplete retrieval of identified research, reporting bias). *Comment on the validity of the assumptions, such as transitivity and consistency. Comment on any concerns regarding network geometry (e.g., avoidance of certain comparisons).* | NA |
| Conclusions | 26 | Provide a general interpretation of the results in the context of other evidence, and implications for future research. | NA |
| **FUNDING** | | | |
| Funding | 27 | Describe sources of funding for the systematic review and other support (e.g., supply of data); role of funders for the systematic review. This should also include information regarding whether funding has been received from manufacturers of treatments in the network and/or whether some of the authors are content experts with professional conflicts of interest that could affect use of treatments in the network. | 18 |

PICOS = population, intervention, comparators, outcomes, study design.
* Text in italics indicateS wording specific to reporting of network meta-analyses that has been added to guidance from the PRISMA statement.
† Authors may wish to plan for use of appendices to present all relevant information in full detail for items in this section.
