## Supplementary File 3. Medline search string for "Relative effectiveness of non-surgical interventions for pain management in knee osteoarthritis: a protocol for a component network meta-analysis of randomised controlled trials"

### Medline (OVID) search string

1 Randomized controlled trial.pt.

2 Controlled clinical trial.pt.

3 randomi?ed.ab.

4 placebo.ab.

5 drug therapy.fs.

6 randomly.ab.

7 trial.ab.

8 groups.ab.

9 or/1-8

10 exp animals/ not humans.sh.

11 9 not 10

12 exp osteoarthritis/

13 (osteoarthriti* or osteo-arthriti* or degenerative joint disease or arthroses or arthrosis or osteoarthros* or coxarthrosis).tw.

14 (degenerative adj2 arthritis).tw.

15 (knee adj4 OA).tw.

16 or/12-15

17 exp pain management/

18 exp mind-body therapies/

19 exp exercise/

20 exp exercise therapy/

21 exp sports/

22 ((strength$ or isometric$ or isotonic$ or isokinetic$ or aerobic$ or endurance or weight$) adj2 (exercis$ or train$)).ti,ab.

23 (resistance training or weight training or physiotherapy or mind?body or tai?ji or tai?chi or taiji or yoga or mind?body or exercise or sport or running or jogging or treadmill or swimming or walking or cycling or rowing or physical activity or physical conditioning or aquarobics or pilates or physical fitness).ti,ab.

24 or/18-23

25 exp patient education as topic/

26 exp *health education/

27 self care/ or (self?care or self?help or self?manage*).ti,ab.

28 ((health or patient$) adj2 (educat$ or information)).tw.

29 or/25-28

30 exp Transcutaneous Electric Nerve Stimulation/

31 Transcutaneous adj4 Stimulation or TENS

32 (electric$ adj (nerve or therapy)).tw.

33 Electromagnetic Fields/
34 electromagnetic$.ti,ab.
35 exp Electric Stimulation Therapy/
36 (electric$ adj3 stimulat$).tw.
37 (alternat$ adj3 electric$).tw.
38 exp ultrasonography/
39 exp Ultrasonic Therapy/
40 us.fs.
41 (ultrasound$ or ultrasonic$).tw.
42 short wave therapy.tw.
43 ultrasonograph$.tw.
44 heat/tu
45 (heat or hot or ice).tw.
46 cryotherapy.sh,tw.
47 (vapocoolant or phonophoresis).tw.
48 exp hyperthermia, induced/
49 (hypertherm$ or thermotherapy).tw.
50 (fluidotherapy or compression).tw.
51 or/30-50
52 24 or 29 or 51
53 exp analgesics/
54 exp acetaminophen/
55 (acetaminophen or acamol or acephen or acetaco or acetamidophenol or acetaminophen or acetominophen or algotropyl or anacin 3 or anacin-3 or anacin3 or datril or hydroxyacetanilide or "n-(4-hydroxyphenyl)acetanilide" or n-acetyl-p-aminophenol or panadol or paracetamol or tylenol or p-acetamidophenol or p-hydroxyacetanilide).ti,ab.
56 or/53-55
57 exp Anti-Inflammatory Agents, Non-Steroidal/
58 aspirin.mp. or exp Aspirin/
59 etodolac.mp. or exp Etodolac/
60 diclofenac.mp. or exp Diclofenac/
61 sulindac.mp. or exp Sulindac/
62 (indometacin or indomethacin).mp. or exp Indomethacin/
63 piroxicam.mp. or exp Piroxicam/
64 fenoprofen.mp. or exp Fenoprofen/
65 flurbiprofen.mp. or exp Flurbiprofen/
66 ibuprofen.mp. or exp Ibuprofen/
67 ketoprofen.mp. or exp Ketoprofen/
68 naproxen.mp. or exp Naproxen/
69 diflunisal.mp. or exp Diflunisal/

70 metamizol.mp. or exp Dipyrone/

71 phenylbutazone.mp. or exp Phenylbutazone/

72 phenazone.mp. or exp Antipyrine/

73 exp cyclooxygenase inhibitors/ or exp cyclooxygenase 2 inhibitors/

74 exp Meclofenamic Acid/

75 tolmetin.mp. or exp Tolmetin/

76 (nsaids or non?steroidal anti?inflammat$ or acetylsalicyl$ or carbasalate calcium or aceclofenac or alclofenac or meloxicam or dexibuprofen or dexketoprofen or tiapro$ or propyphenazone or celecoxib or etoricoxib or nabumeton or parecoxib or ((cyclooxygenase or cyclo-oxygenase) adj3 inhibitor*) or rofecoxib or valdecoxib or lumiracoxib or vioxx or celebrex or bextra or prexige or arcoxia or floctafenine or meclofenamate or oxaprozin or tenoxicam).mp.

77 or/57-76

78 (bufexamac OR bufexine OR calmaderm OR ekzemase OR diclofenac OR solaraze OR pennsaid OR voltarol OR emugel OR voltarene OR voltarol OR optha OR voltaren OR etofenamate OR afrolate OR algesalona OR bayro OR deiron OR etofen OR flexium OR flogoprofen OR rheuma-gel OR rheumon OR traumalix OR traumon OR zenavan OR felbinac OR dolinac OR flexfree OR napageln OR target OR traxam OR fentiazac OR domureuma OR fentiazaco OR norvedan OR riscalon OR fepradinol OR dalgen OR flexidol OR cocresol OR rangozona OR reuflodol OR pinazone OR zepelin OR flufenamic OR dignodolin OR rheuma OR lindofluid OR sastridex OR lunoxaprofen OR priaxim OR flubiprofen OR fenomel OR ocufen OR ocuflur OR tulip OR ibuprofen OR cuprofen OR "deep relief" OR fenbid OR ibu?cream OR ibugel OR ibuleve OR ibumousse OR ibuspray OR "nurofen gel" OR proflex OR motrin OR advil OR radian OR ralgex OR ibutop OR indomethacin OR indocin OR indospray OR isonixin OR nixyn OR ketoprofen OR tiloket OR oruvail OR powergel OR solpaflex OR ketorolac OR acular OR trometamol OR meclofenamic OR naproxen OR naprosyn OR niflumic OR actol OR flunir OR niflactol topico OR niflugel OR nifluril OR oxyphenbutazone OR californit OR diflamil OR otone OR tanderil OR piketoprofen OR calmatel OR triparsean OR piroxicam OR feldene OR pranoprofen OR oftalar OR pranox OR suxibuzone OR danilon OR flamilon OR ufenamate OR fenazol OR flector OR benzydamine).mp.

79 or/77-78

80 exp Administration, Topical/ or (topical* OR cutaneous OR dermal OR transcutaneous OR transdermal OR percutaneous OR skin OR massage OR embrocation OR gel OR ointment OR aerosol OR cream OR creme OR lotion OR mouse OR foam OR liniment OR spray OR rub OR balm OR salve OR emulsion OR oil OR patch OR plaster).mp.

81 79 AND 80

82 capsaicin.mp. AND 80

83 exp analgesics, opioid/ or (alfentanil or alphaprodine or buprenorphine or butorphanol or codeine or dextromoramide or dextropropoxyphene or dihydromorphine or diphenoxylate or

ethylketocyclazocine or ethylmorphine or etorphine or fentanyl or hydrocodone or hydromorphone or levorphanol or meperidine or meptazinol or methadone or methadyl acetate or morphine or nalbuphine or opiate alkaloids or opium or oxycodone or oxymorphone or pentazocine or phenazocine or phenoperidine or pirinitramide or promedol or remifentanil or sufentanil or tapentadol or tilidine or tramadol).mp.

84 (cymbalta or duloxetine).mp.

85 exp hyaluronic acid/ or viscosupplement/ or viscosupplementation/ or (hyaluronic or hyaluronate* or hyaluronan*).mp.

86 (adant or arthrease or arthrum h or artz or artzal or biohy or durolane or etapharm or euflexxa or fermathron or go-on or healon or healonid or hyalflex or hyalgan or hyalurons or hylan* or hylartil or hylectin or hyruan or nasha or neovisc or nrd-101 or nuflexxa or orthovisc or ostenil or polireumin or polyreumin or replasyn or slm-10 or supartz or suplasyn or suvenyl or synject or synocrom or synvisc).mp.

87 or/85-86

88 *Adrenal Cortex Hormones/ or *17-Hydroxycorticosteroids/ or *11-Hydroxycorticosteroids/ or *Hydroxycorticosteroids/ or *Ketosteroids/ or *17-Ketosteroids/ or *Androstenedione/ or *Prednisolone/ or *Glucocorticoids/ or *Triamcinolone Acetonide/ or *Hydrocortisone/ or *cortisone/

89 (adrenal cortex hormone* or adrenal cortical hormone* or adrenal steroid* or adrenocortical hormone* or adrenocortical steroid* or adrenocorticalsteroid* or adrenocorticosteroid* or cortical steroid* or cortico-steroid* or corticoid* or corticosteroid* or dermocortico-steroid* or dermocorticosteroid* or glucocortic* or hydroxycorticosteroid* or ketosteroid* or androstenedion* or steroid or triamcinolone hexacetonide or hydrocortison* or prednisolone or Prednison* or cortison* or Pregnadiene*).mp.

90 or/88-89

91 (intraartic* or intra-artic* or inject* or infiltration* or infiltrating).mp.

92 and/90,91

93 56 or 77 or 81 or 82 or 83 or 84 or 87 or 87 or 92

94 17 or 52 or 93

95 11 AND 16 AND 94
